# Comparison of antibody responses to SARS-CoV-2 variants in Australian children

**DOI:** 10.1101/2022.08.12.22278705

**Authors:** Zheng Quan Toh, Nadia Mazarakis, Jill Nguyen, Rachel A. Higgins, Jeremy Anderson, Lien Anh Ha Do, David P. Burgner, Nigel Curtis, Andrew C. Steer, Kim Mulholland, Nigel W. Crawford, Shidan Tosif, Paul V Licciardi

## Abstract

There is limited understanding of antibody responses in children across different SARS-CoV-2 variants. As part of an ongoing household cohort study, we assessed the antibody response among unvaccinated children infected with Wuhan, Delta or Omicron variants, as well as vaccinated children with breakthrough Omicron infection, using a SARS-CoV-2 S1-specific IgG assay and surrogate virus neutralisation test (sVNT). Most children infected with Delta (100%, 35/35) or Omicron (81.3%, 13/16) variants seroconverted by one month following infection. In contrast, 37.5% (21/56) children infected with Wuhan seroconverted, as previously reported. However, Omicron-infected children (GMC 46.4 BAU/ml; sVNT % inhibition: 16.3%) mounted a significantly lower antibody response than Delta (435.5 BAU/mL, sVNT=76.9%) or Wuhan (359.0 BAU/mL, sVNT=74.0%). Vaccinated children with breakthrough Omicron infection mounted the highest antibody response (2856 BAU/mL, sVNT=96.5%). Our findings suggest that despite a high seroconversion rate, Omicron infection in children results in lower antibody levels and function compared with Wuhan or Delta infection or with vaccinated children with breakthrough Omicron infection. Our data have important implications for public health measures and vaccination strategies to protect children.

## Main text

Children have been less likely to be infected and develop severe disease by the original SARS-CoV-2 (Wuhan) strain compared to adults^1-3^. However, the combination of increased transmissibility of SARS-CoV-2 Delta and Omicron variants, increased population movement due to easing of COVID-19 restrictions, and a higher vaccination rate in adults compared with children have resulted in rising COVID-19 cases among children^4,5^. Despite this, SARS-CoV-2 infections in children are mostly mild or asymptomatic.

In Australia, there have been three epidemic waves of COVID-19, caused respectively by the original Wuhan strain (first infection documented in March 2020), the Delta variant (May 2021) and the Omicron variant (November 2021)^6^. Australian children aged 5-11 years were eligible for COVID-19 vaccination from December 2021, with uptake of two doses estimated at 40.6% as of 7^th^ August 2022^5^.

The Omicron variant BA.1/BA.2 is associated with reduced clinical severity and risk of hospitalization compared to the Delta variant in both children and adults^7-10^. Adults mount strong Omicron-specific humoral responses^11^, but limited data are available in children. We previously reported that only 37.5% of children infected with the Wuhan strain seroconverted compared with 76.2% of adults^12^. It is unknown if a similar pattern occurs following Delta or Omicron infection in children. We therefore aimed to compare seroconversion rates and antibody responses in children across the three SARS-CoV-2 waves in Melbourne, Australia.

In this household cohort study, suspected SARS-CoV-2 cases and household members of suspected cases were tested by RT-PCR on nasopharyngeal (NP) swabs at The Royal Children’s Hospital or by nasal rapid antigen test (RAT) at home. Confirmed SARS-CoV-2 cases and household members were invited to participate. Blood samples were collected approximately one month following PCR/RAT diagnosis. Written informed consent and assent were obtained from adults/parents and children, respectively. The study was conducted with the approval of the Royal Children’s Hospital Human Research Ethics Committee (HREC): HREC/63666/RCHM-2019. A SARS-CoV-2 S1 in-house ELISA and surrogate virus neutralization test (sVNT) were used to measure IgG and neutralising antibody responses using Wuhan and variant-specific S1 antigens. Detailed methodology, including description of epidemic waves, ELISA and sVNT assays and statistical analyses, are provided in the Supplementary material.

Between March 2020 and July 2022, a total of 580 adults and children were enrolled. Participants aged between 6 months to 17 years with COVID-19 confirmed by SARS-CoV-2 PCR or RAT on nasopharyngeal swab who had not received any COVID-19 vaccine were included in this analysis (n=107). We also included children who had received at least one dose of a COVID-19 vaccine but experienced a breakthrough Omicron infection (n=24, extended data, Figure 1 and Table 1). Unvaccinated children infected with Omicron were younger than children infected with Wuhan or Delta variant

**Figure 1:**
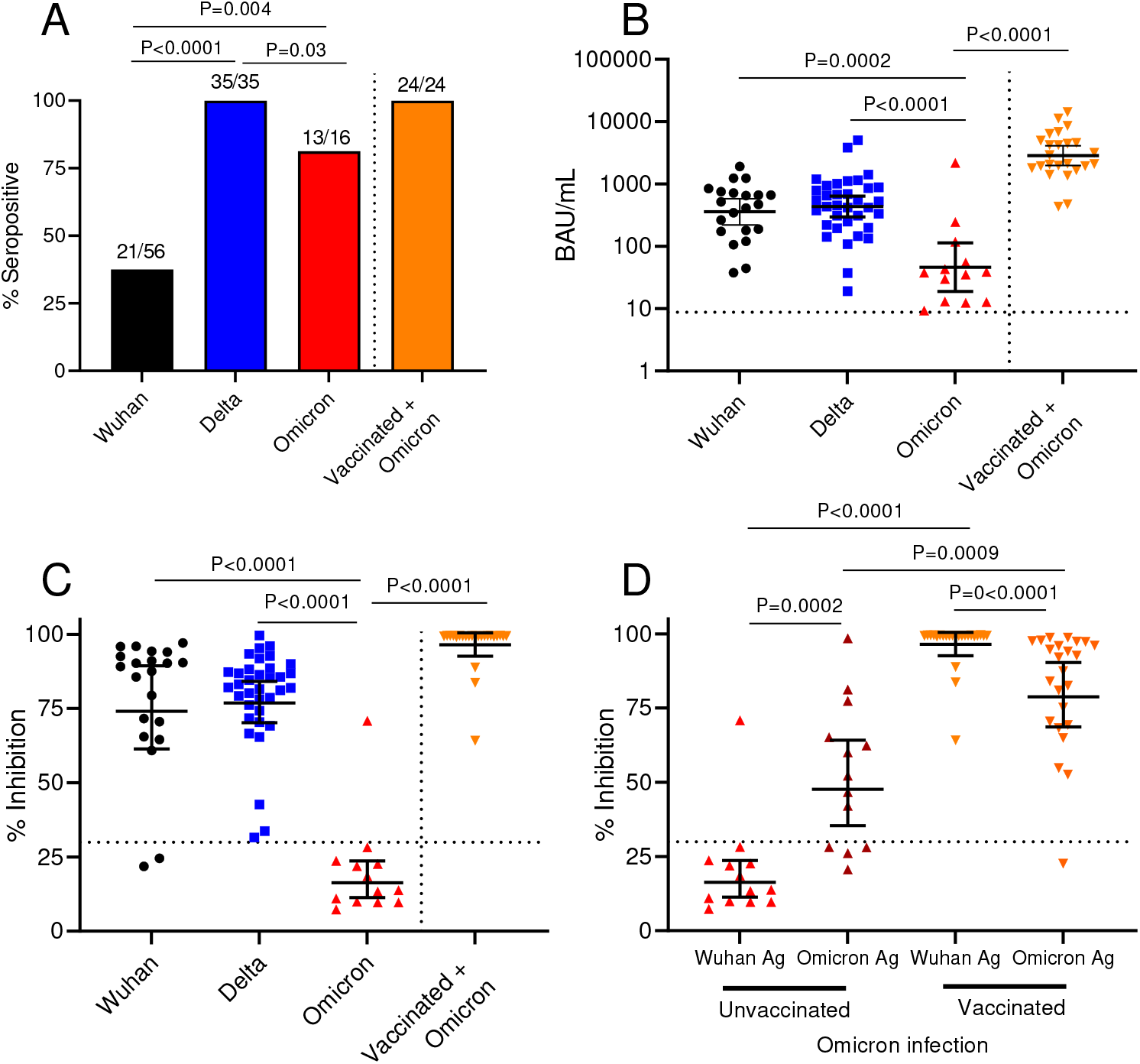
SARS-CoV-2 antibody responses in children infected with Wuhan, Delta or Omicron variant and children previously vaccinated and then infected with Omicron. (A) SARS-CoV-2 Immunoglobulin G (IgG) Seropositivity Rate at Day 36 post-infection. Among children who seroconverted at Day 36 post-infection, (B) SARS-CoV-2 IgG concentrations, (C) SARS-CoV-2 neutralising antibodies as measured by surrogate virus neutralisation test. (D) SARS-CoV-2 neutralising antibodies as measured by surrogate virus neutralisation test using Wuhan- or Omicron-specific antigen. Dotted lines indicate seropositivity cut-off. Each data point represents an individual participant. Data presented as geometric mean concentrations ± 95% confidence intervals. BAU, binding antibody units.

Among the 56 children infected with the Wuhan strain, 21 (37.5%) seroconverted by day 36^12^, while all 35 children infected with Delta (P<0.0001), and 13 of 16 (81%) infected with Omicron (P=0.004) variants seroconverted (Figure 1A). Two out of three children infected with Omicron were seronegative based on testing using the Wuhan S1 antigen but were seropositive when tested using the Omicron S1 antigen.

Among children who seroconverted, S1-specific IgG concentrations to Omicron infection (GMC: 46.4 BAU/ml, 95% CI: 18.9, 113.8) were 7.7- and 9.4-fold lower than Wuhan (GMC: 359 BAU/ml, 95% CI: 221.2, 582.5, P=0.0002) or Delta infections (GMC: 435.5 BAU/ml, 95% CI: 296.9, 638.9, P<0.0001), respectively (Figure 1B). Vaccinated children with Omicron breakthrough infection had 61.6-fold higher IgG GMC than unvaccinated Omicron-infected children. Similarly, lower neutralizing antibody responses based on the Wuhan-antigen-based sVNT were observed among unvaccinated Omicron-infected children (only one out of the 13 children who seroconverted had a positive neutralising antibody response) compared with children who seroconverted following Wuhan or Delta infection or who were previously vaccinated (19 of 21, 35 of 35 and 24 of 24 had high neutralising antibody responses respectively, P<0.0001 for all comparisons, Figure 1C). This lower response to Omicron was not associated with younger age when we stratified the analysis based on 5-17 years and <5 years for Wuhan and Delta (extended data, Figure 2). When we used variant-specific S1 antigens, we observed higher IgG concentrations for Omicron but not Delta (extended data, Figure 3). Omicron-specific neutralizing antibody responses were detected in nine out of 13 children when measured using the Omicron-specific sVNT for the Omicron cohort (geometric mean % inhibition: 47.7%, 95% CI 35.4%, 64.2%), but the responses were still significantly lower compared with vaccinated children with Omicron breakthrough 78.8% (95% CI 68.7%, 90.4%, P=0.009, Figure 1D). Children infected with Wuhan or Omicron, but not Delta, had significantly lower IgG concentrations than adults (2.1-fold and 89.1-fold, respectively), although the number of unvaccinated adults in the Omicron cohort was small due to high vaccine coverage among adults in the cohort (extended data, Figure 4). The neutralizing antibody response against Omicron was consistent with the IgG response (extended data, Figure 4).

Our findings indicate that almost all children seroconverted following Omicron infection. However, these children mounted a lower antibody response compared to the Wuhan strain or the Delta variant, which contrasts with observations in adults^11^. How this relates to protection against re-infection and disease is unknown. The role of other immune factors (i.e. cellular and mucosal immunity) in protection against Omicron re-infection, particularly BA.4/5 subvariants among unvaccinated and vaccinated children remains to be determined. Vaccinated children with breakthrough Omicron infection mounted the highest antibody responses in our study, suggesting that vaccination may help overcome the low antibody response following Omicron variant infection in children. We also found that the antibody response against Omicron may be substantially underestimated when measured using Wuhan antigen, with implications for seroprevalence studies and estimating protective antibody titres against Omicron infection. On 3^rd^ August 2022, Australia authorities recommended the use of the mRNA-1273 (Moderna^®^) vaccine for high-risk children aged from 6 months to < 5 years^13^. Ongoing serological studies to monitor immunogenicity against SARS-CoV-2 variants of concern remains crucial. Furthermore, new formulations of vaccine(s) are under review by international regulators, including bivalent Ancestral/Omicron vaccines^14^, and our household cohort study will continue to monitor immunological responses when these new-generation vaccines become available to the paediatric population. Limitations of the study include the small sample size, particularly for the Omicron cohort and the young age group (<5 years) in the Omicron cohort, since most children in our cohort were ≥5 years and had received at least one dose of COVID-19 vaccine. We also did not have data on viral loads and information on the infecting strain. However, we believe our assumptions about the infecting variant in each cohort were correct based on available local epidemiological and sequence data^6^.

In summary, the immune response to SARS-CoV-2 infection among children varies between the variants. While most unvaccinated children infected with Delta or Omicron seroconvert, antibody responses to Omicron were much lower. This might lead to an increased risk of repeated re-infection of children, which would in-turn have serious implications for their long-term health. The much stronger antibody responses observed among vaccinated children supports vaccination of children to induce greater protective immunity.

## Data Availability

All data produced in the present work are contained in the manuscript

## Acknowledgments

We thank the study participants and families for their involvement in this study. We also acknowledge the SAEFVIC Research Team (Alissa McMinn, Hayley Giuliano, Kate Hession, Belle Overmars, Chelsea Bartel). We also acknowledge the Murdoch Children’s Research Institute (MCRI) Biobanking service for their help in processing the samples. Funding for the recruitment of participants was provided by the Royal Children’s Hospital Foundation and the MCRI Infection and Immunity Theme, grants from the DHB Foundation and the Victorian Department of Jobs, Precincts and Regions. This work is supported by Victorian Government’s Medical Research Operational Infrastructure Support Program. MCRI. PVL is supported by NHMRC Career Development Fellowship. DB, ACS and NC are supported by NHMRC Investigator Grants. ACS is supported by a Viertel Senior Medical Research Fellowship. ST is supported by a MCRI Clinician Scientist Fellowship. ZQT is supported by an MCRI Early Career Fellowship. NWC received funding from the National Institute of Health for influenza and COVID-19 research. All other authors reported no conflicts of interests.

## Figure legends

**Extended data, Figure 1: Overview of the cohort by age and their date of SARS-CoV-2 infection**. Each dot represents a child. N=131.

**Extended data, Figure 2: SARS-CoV-2 antibody responses in children infected with Wuhan, Delta or Omicron strain stratified by age**. Each data point represents an individual participant. Data presented as geometric mean concentrations ± 95% confidence intervals. BAU, binding antibody units.

**Extended data, Figure 3: SARS-CoV-2 antibody responses in children infected with Wuhan, Delta or Omicron strain measured using Wuhan or variant-specific S1 antigen (Ag)**. SARS-CoV-2 IgG concentrations measured by ELISA assay. Dotted lines indicate seropositivity cut-off. Each data point represents an individual participant. Data presented as geometric mean concentrations ± 95% confidence intervals. BAU, binding antibody units.

**Extended data, Figure 4: Comparison of SARS-CoV-2 antibody responses between children and adults**. (A) SARS-CoV-2 IgG concentration in children (C) or adults (A) infected with Wuhan, Delta or Omicron strain. (B) Neutralising antibodies of unvaccinated children or adults infected with Omicron measured by SARS-CoV-2 surrogate virus neutralisation assay using Wuhan or Omicron antigen (Ag). Each data point represents an individual participant. Dotted lines indicate seropositivity cut-off. Data presented as geometric mean concentrations ± 95% confidence intervals. BAU, binding antibody units.

