## Extended data for "Comparison of antibody responses to SARS-CoV-2 variants in Australian children"

### Slide 1
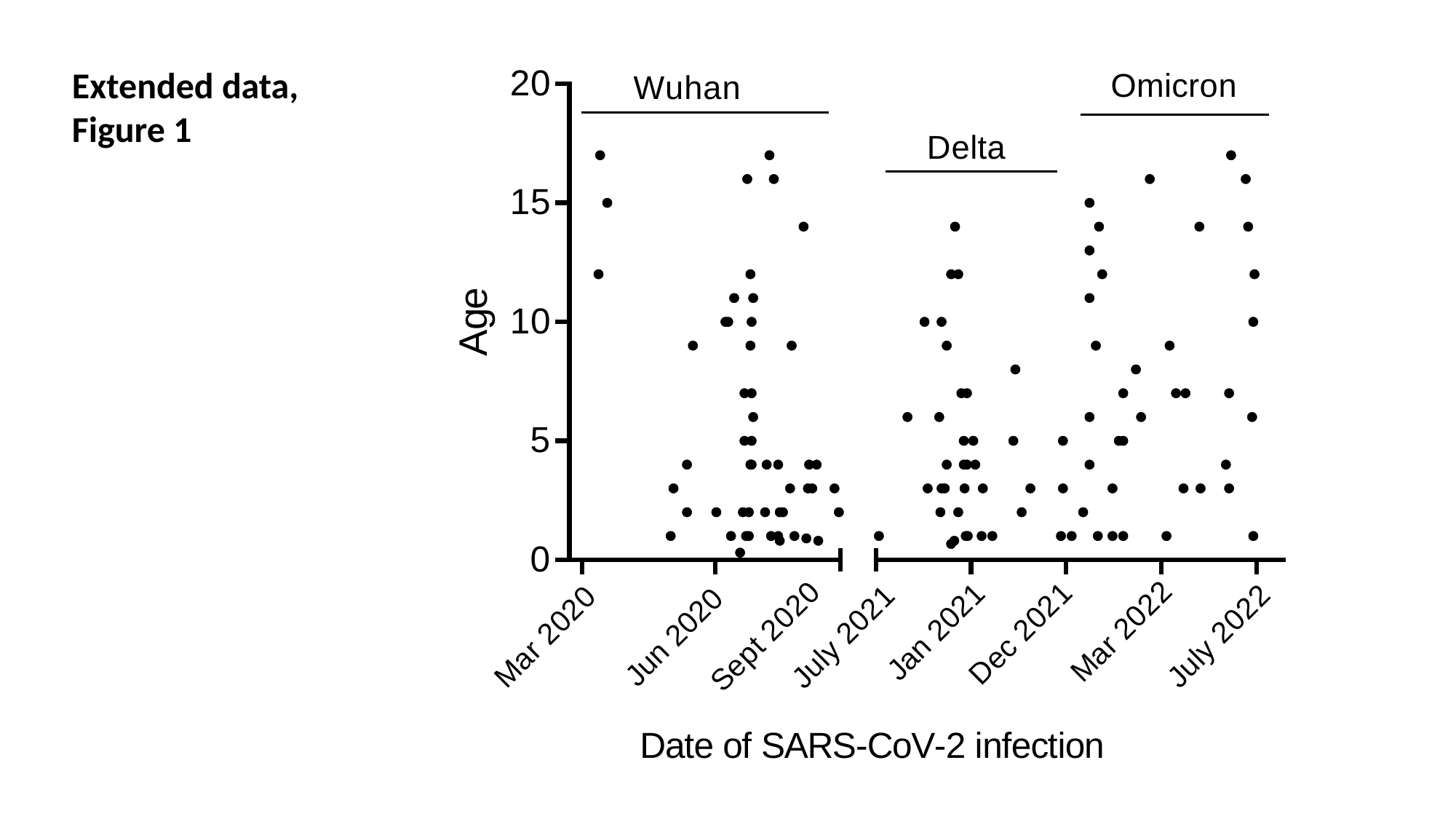

Extended data, Figure 1

### Slide 2
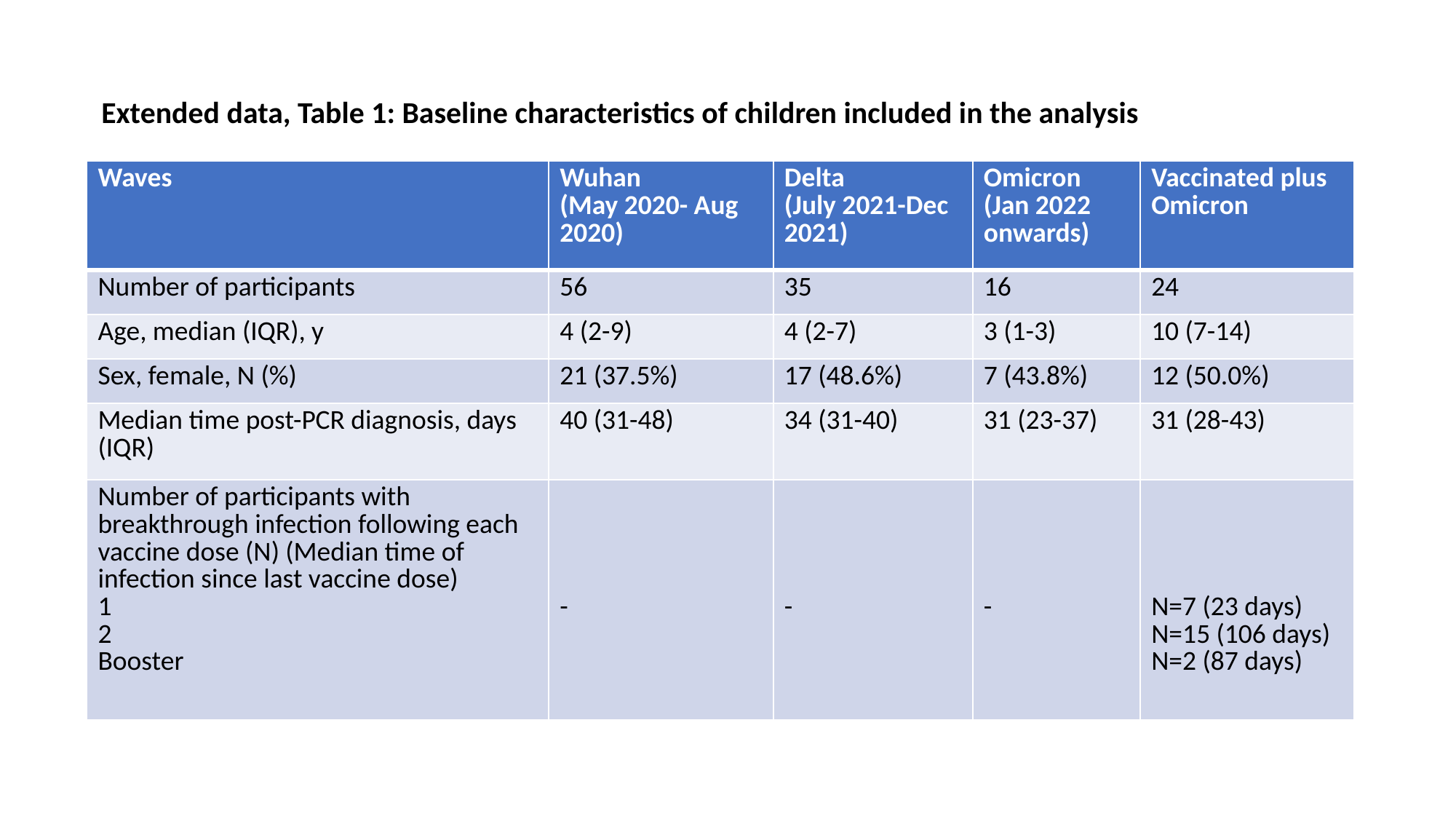

Extended data, Table 1: Baseline characteristics of children included in the analysis
| Waves | Wuhan (May 2020- Aug 2020) | Delta (July 2021-Dec 2021) | Omicron (Jan 2022 onwards) | Vaccinated plus Omicron |
| --- | --- | --- | --- | --- |
| Number of participants | 56 | 35 | 16 | 24 |
| Age, median (IQR), y | 4 (2-9) | 4 (2-7) | 3 (1-3) | 10 (7-14) |
| Sex, female, N (%) | 21 (37.5%) | 17 (48.6%) | 7 (43.8%) | 12 (50.0%) |
| Median time post-PCR diagnosis, days (IQR) | 40 (31-48) | 34 (31-40) | 31 (23-37) | 31 (28-43) |
| Number of participants with breakthrough infection following each vaccine dose (N) (Median time of infection since last vaccine dose) 1 2 Booster | - | - | - | N=7 (23 days) N=15 (106 days) N=2 (87 days) |

### Slide 3
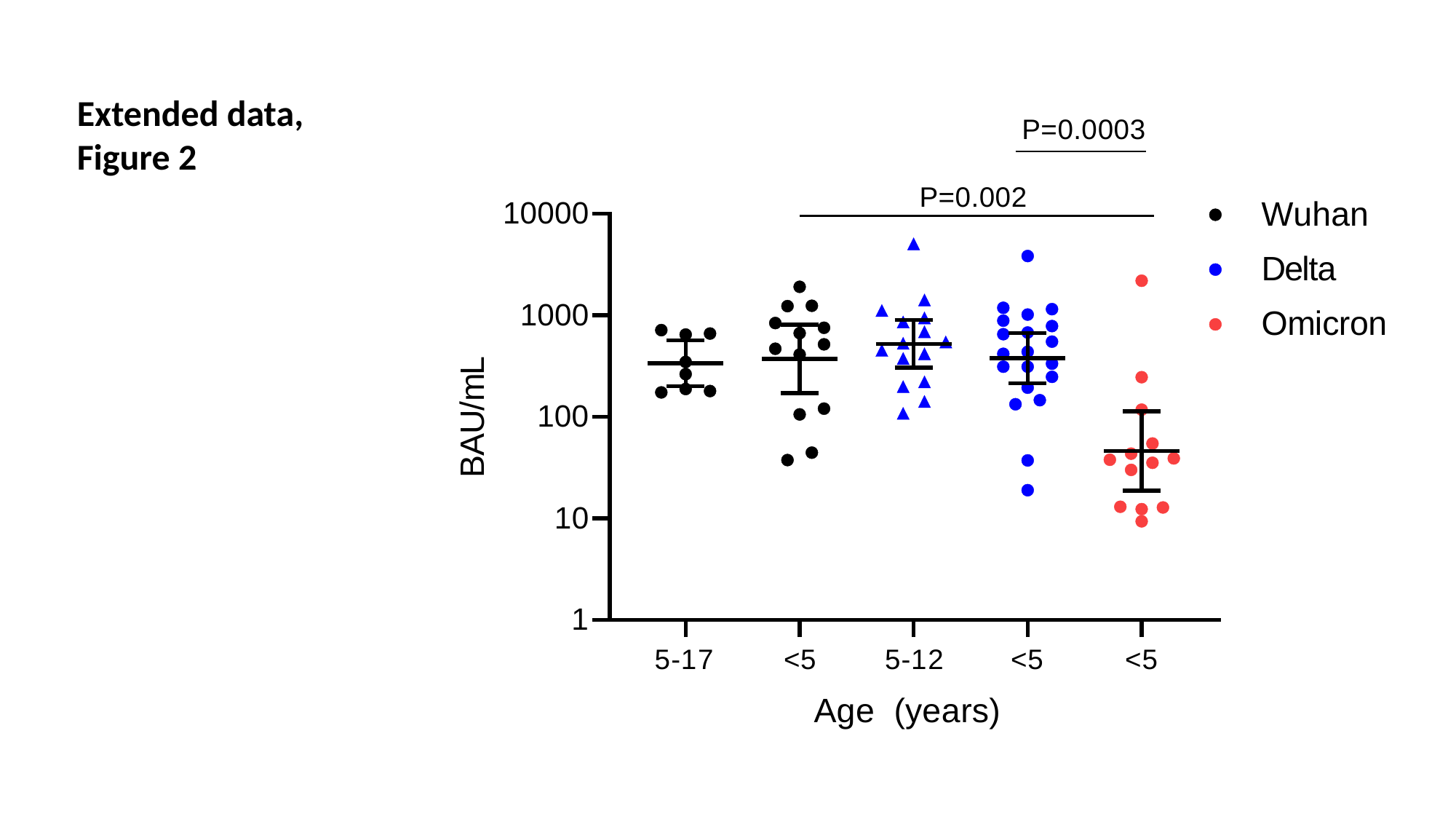

Extended data, Figure 2

### Slide 4
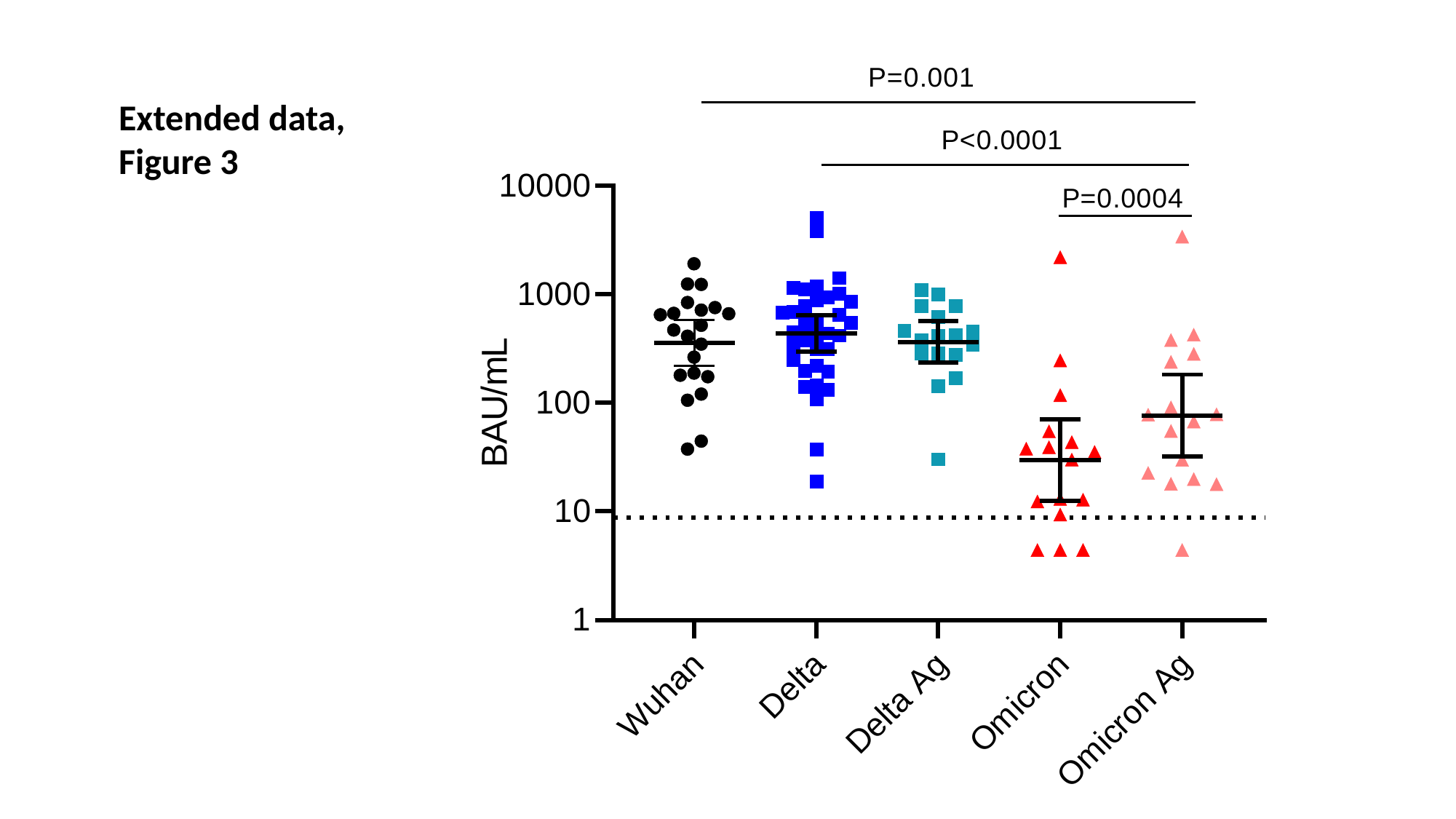

Extended data, Figure 3

### Slide 5
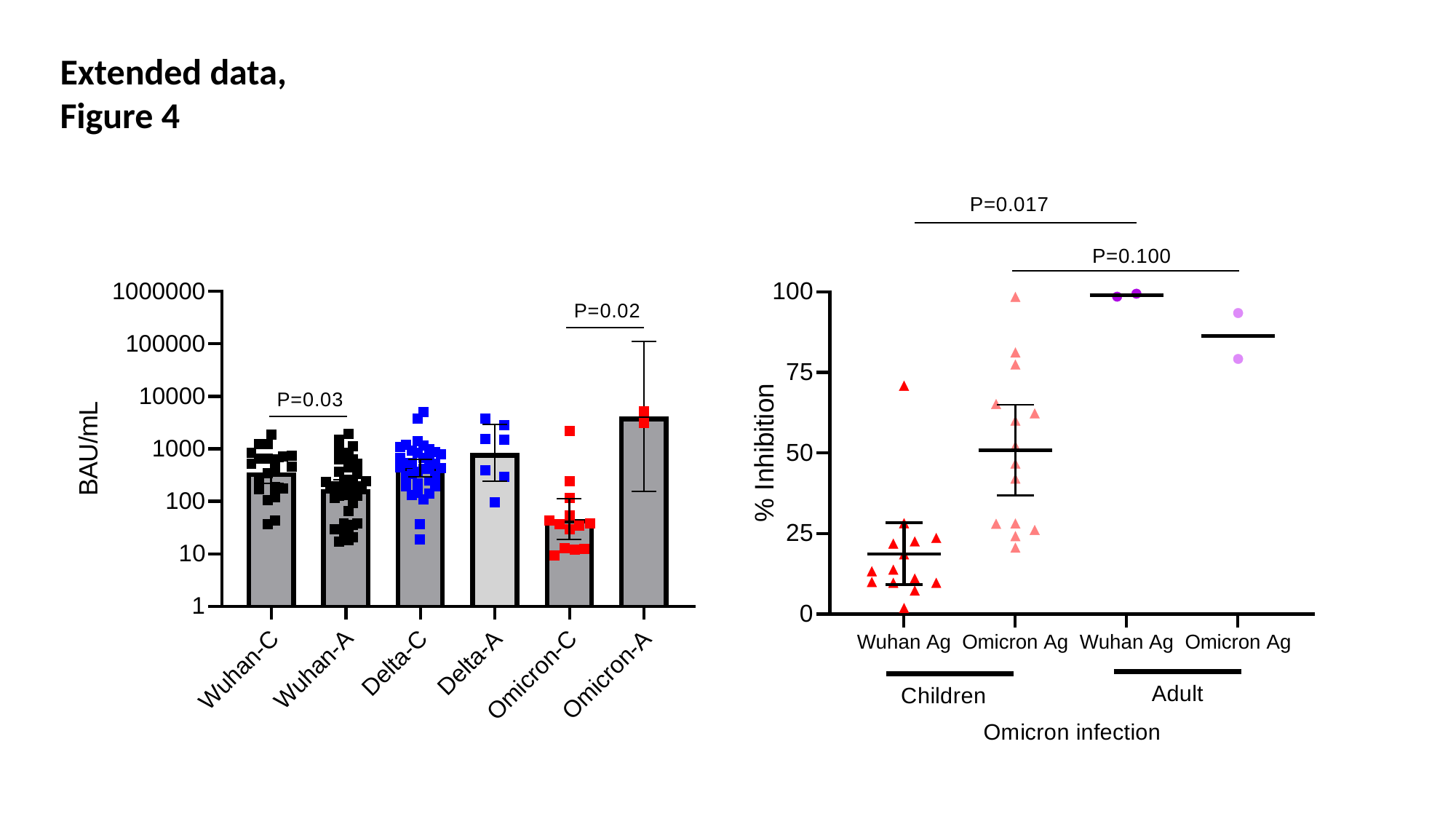

Extended data, Figure 4
