## Supplementary methods for "Comparison of antibody responses to SARS-CoV-2 variants in Australian children"

**Supplementary Information**

**Methods**

*Epidemic waves*

As of July 2022, Australia has had three epidemic waves of COVID-19 caused by the original (ancestral) Wuhan strain (first infection documented in March 2020), Delta (May 2021) and Omicron (November 2021) variants. In Victoria, Australia, the Delta and Omicron variant was first detected in late May 2021 and early December 2021, respectively. The Delta cohort was recruited between July 2021 and November 2021, while the Omicron cohort was recruited between January 2022-July 2022 based on data from Nextstrain.org

*SARS-CoV-2 ELISA method*

Details of the ELISA method has been previously published^1,2^. The ELISA method was previously validated against two commercial assays (Diasorin LIAISON^®^ and Wantai) and a SARS-CoV-2 micro neutralisation assay ^1,2^. The variant-specific S1 antigens (Delta and Omicron) were sourced from Sino Biological (Sino Biological Inc., China <https://www.sinobiological.com>). Seropositive samples were titrated and calculated based on a World Health Organization SARS-CoV-2 pooled serum standard (NIBSC code: 20/130, National Institute of Biological Standards and Controls, UK). Results are reported in binding antibody units per milliliter (BAU/mL). The cutoff for seropositivity was 8.82 BAU/mL based on pre-pandemic samples, whereas seronegative samples were given half of the seropositive cutoff value.

*SARS-CoV-2 surrogate viral neutralisation test (sVNT)*

SARS-CoV-2 surrogate virus neutralisation test (sVNT) was conducted according to the manufacturer’s instructions (GenScript, New Jersey, USA). For Omicron sVNT, an Omicron specific HRP-conjugated RBD was used instead of the Wuhan HRP-RBD, and the Omicron-specific neutralizing antibody standard was used as the positive control. The results were reported as a percentage (%) of inhibition by neutralising antibodies in relation to the negative control. Results <30% inhibition are considered negative, while ≥30% inhibition are considered positive for neutralising antibodies.

*Statistical Analysis*

Proportion of participants who seroconverted between different variants were compared using Fisher’s Exact test. Antibody concentrations and sVNT neutralizing antibodies were reported as geometric mean concentration (GMC) and percent inhibition, respectively. The S1-specific IgG antibody concentrations (BAU/ml) and neutralizing antibodies (%) between children from different SARS-CoV-2 waves, between unvaccinated and vaccinated, as well as between different age groups were compared using Mann-Whitney U test. For comparison of variant-specific antibody responses, Wilcoxon-signed rank test were used. All analyses were performed with GraphPad Prism version 9.0 (GraphPad Software, https://www.graphpad.com). A p<0.05 was considered significant.
